# Multi-Agent AI for Chest Radiography: A Sequential Segmentation and LLM-Driven Consultative Tool for Medical Training

**DOI:** 10.64898/2026.05.29.26354432

**Authors:** Furkan Kurt, Abdulhamit Subasi

## Abstract

**Background:** Traditional diagnostic models lack explainability, while multimodal language models prone to hallucination remain unsafe for medical education. An interactive, risk-free artificial intelligence framework is required to serve as a reliable clinical mentor for radiology trainees.

**Methods:** We propose a multi-agent architecture decoupling deterministic image analysis from generative consultation. Specialized computer vision models perform anatomical localization and pathological segmentation. These quantitative outputs are synthesized into a structured payload, which grounds a locally hosted large language model (LLaVA 7B) using strict prompt guardrails and prerequisite protocols.

**Results:** The system effectively eliminates visual hallucinations by intercepting unanchored queries. The artificial intelligence tutor successfully contextualizes spatial anomalies and baseline metrics, generating accurate conversational explanations and formally structured radiology reports while strictly enforcing medical safety disclaimers.

**Discussion and Conclusion:** By anchoring language generation exclusively to verified algorithmic realities, this framework transforms opaque diagnostic models into safe, interactive educational simulators. This establishes a highly reliable paradigm for integrating explainable artificial intelligence into medical training.

## 1. Introduction

Mastering chest X-ray interpretation is a critical but highly challenging competency in medical education, often characterized by a steep learning curve and significant diagnostic variability among early-career clinicians [1]. While advanced deep learning techniques have demonstrated remarkable clinical accuracy, achieving upwards of 98% accuracy in complex image classification tasks [2], their integration into medical training remains fundamentally limited. Traditional Computer-Aided Diagnosis systems operate predominantly as opaque models. They generate end-stage probability scores or simple binary classifications but fail to articulate the underlying visual and clinical reasoning required for a student to learn. Consequently, these static diagnostic tools are inadequate for medical trainees who require step-by-step guidance to develop independent diagnostic logic. Recent literature emphasizes an urgent need within medical education to transition from passive diagnostic aids to interactive, AI-driven intelligent tutoring systems [3]. Interactive educational frameworks have proven highly effective in increasing learner confidence, providing immediate objective feedback, and reducing the fear of making clinical errors in a risk-free setting [1], [4].

Large Language Models (LLMs) present a promising avenue for creating these interactive clinical simulators, yet their deployment in radiology is severely hindered by the phenomenon of medical hallucinations. Recent evaluations of multimodal models reveal a high susceptibility to generating confident but incorrect outputs in medical imaging tasks [5]. These errors manifest frequently in image interpretation, with models demonstrating false positives, hallucinated absences of actual diseases, and clinically implausible anatomical descriptions [5]. Because current language models can hallucinate at a baseline rate of 8 to 15 percent in medical contexts, their unconstrained use limits their clinical trustworthiness and poses a risk of misleading medical trainees [6].

To address this critical gap and mitigate the risk of fabricated medical data, researchers are increasingly exploring agentic AI approaches. Dividing complex clinical tasks among specialized, role-based agents has been shown to effectively reduce hallucinations by anchoring the language model to verify clinical data [6]. Building upon this paradigm, this paper introduces a novel, sequential multi-agent artificial intelligence pipeline design specifically for chest radiography education. Rather than relying on a single multimodal model to blindly interpret an image, our system functions as an explainable educational mentor. It sequentially combines precise pixel-level computer vision segmentation models with a strictly guard railed, locally executed LLM. By anchoring the conversational interface to a structured diagnostic ground truth derived from the vision agents, the system eliminates the risk of medical hallucinations while providing interactive, step-by-step clinical mentorship.

This study aims to evaluate the architectural feasibility and educational potential of this system by addressing the following research questions:

- **RQ1:** How can distinct anatomical and pathological segmentation models be integrated into a real-time, multi-agent pipeline to establish a reliable clinical ground truth?
- **RQ2:** How can prompt engineering and prerequisite guardrails effectively constrain Large Language Models to prevent medical hallucinations during consultative dialogues?
- **RQ3:** To what extent can this sequential AI approach successfully simulate an interactive, risk-free clinical mentorship environment for medical trainees?

## 2. Background & Related Work

The shift toward deep learning frameworks like Convolutional Neural Networks (CNNs) and Vision Transformers has greatly improved diagnostic accuracy in chest radiography. These advanced models perform at expert levels, but their complex structures make them opaque and obscure their internal decision-making processes [7]. This lack of transparency creates significant challenges for medical education because high statistical reliability does not automatically translate to clear clinical interpretability [8]. Providing medical trainees with only a probability score fails to teach independent diagnostic reasoning. In response, researchers have turned to Explainable AI techniques, mostly using visual attribution methods like saliency maps and heatmaps to bridge this gap [9].

Despite their popularity, these visual overlays suffer from severe methodological flaws and are often disconnected from the actual mathematical reasoning of the model. These interpretability tools frequently highlight irrelevant image artifacts or medical equipment instead of genuine physiological issues [7], [8], and using more advanced networks can even result in scattered, confusing attention patterns rather than clear explanations [10]. Current explainability methods rely too heavily on gradient-based approaches that miss causal links and ignore important clinical contexts like patient comorbidities [11]. Because simple heatmaps only indicate where a model looks without explaining why it matters clinically, their subjective nature makes them inadequate for training purposes [9]. To truly benefit medical education, systems must evolve past basic visual overlays and integrate precise visual localization with explicit clinical reasoning [7].

Radiology education demands that students quickly master complex anatomy and subtle visual signs of disease, a steep learning curve traditionally managed through direct apprenticeship. Because heavy clinical workloads prevent educators from offering continuous personalized feedback, medical programs are increasingly adopting structured digital interventions to support foundational teaching [12], [13]. Transitioning to active participation is vital for building diagnostic confidence, and interactive adaptive tutorials have proven to be highly efficient and engaging alternatives to standard resources [14]. By incorporating strategies like gamification and simulation, these digital tools serve as powerful complements to face-to-face instruction by turning passive observation into an active cognitive process [12], [13], [15].

Even with their proven success, conventional eLearning platforms still fall short of replicating the dynamic flexibility of a human mentor. Intelligent tutoring systems and artificial intelligence are uniquely positioned to fill this specific apprenticeship gap by offering a safe environment where trainees can test clinical hypotheses and receive immediate feedback without endangering patients [15]. This cycle of guided trial and error is fundamental for cultivating strong diagnostic reasoning. Consequently, developing an AI mentor that merges the broad scalability of digital platforms with the interactive depth of human teaching marks the essential next step in the evolution of medical training.

Integrating language models into healthcare offers great potential, especially for text-based tasks like simplifying radiology reports, where they perform well with very few errors [16]. However, using these models directly for image-based diagnostics introduces severe safety risks. When multimodal models try to process medical images and generate text at the same time, they frequently create false information, commonly known as hallucinations. Studies show that these errors occur in over 70 percent of unassisted visual assessments [17]. These models often miss basic pathological features, invent diseases that are not actually there, and rely too heavily on textual hints rather than genuine visual reasoning [17], [18].

The clinical impact of these errors is highly concerning. Surveys reveal that most clinicians have encountered medical artificial intelligence hallucinations, viewing them as direct threats to patient safety, with many errors capable of causing major harm [19], [20]. Because relying on a single, unconstrained model is too dangerous for medical training, researchers are shifting toward multi-stage, agentic artificial intelligence frameworks. Instead of using one model to do everything, these systems divide complex clinical tasks among multiple specialized models [6]. This separation of tasks, especially utilizing dedicated safety mechanisms, has successfully reduced clinical safety violations from nearly 20 percent to under 3 percent [21].

A critical part of these multi-agent systems is keeping information retrieval strictly separate from text generation. By anchoring the language model to verified data sources through structured retrieval processes, diagnostic accuracy improves significantly while false claims drop [22]. In some specific tasks, grounding the system in verified external data has eliminated hallucinations [6]. These results prove that simply fine-tuning a model is not enough; strong procedural rules and cross-validation are required to keep errors below standard human error rates [19], [20]. Therefore, a safe radiology training simulator must use a sequential pipeline where precise visual algorithms establish clinical facts first, strictly preventing the language model from generating unverified medical claims.

## 3. Methodology

### 3.1. High-Level System Architecture and Orchestration

To address the limitations of opaque diagnostic models and establish an interactive educational environment, the proposed system employs a highly modular, three-tier architecture [Figure 1]. This structural design strictly decouples user interface orchestration, multi-model visual inference, and generative language consultation into distinct operational layers, ensuring both high computational efficiency and strict data privacy.

**Figure 1:**
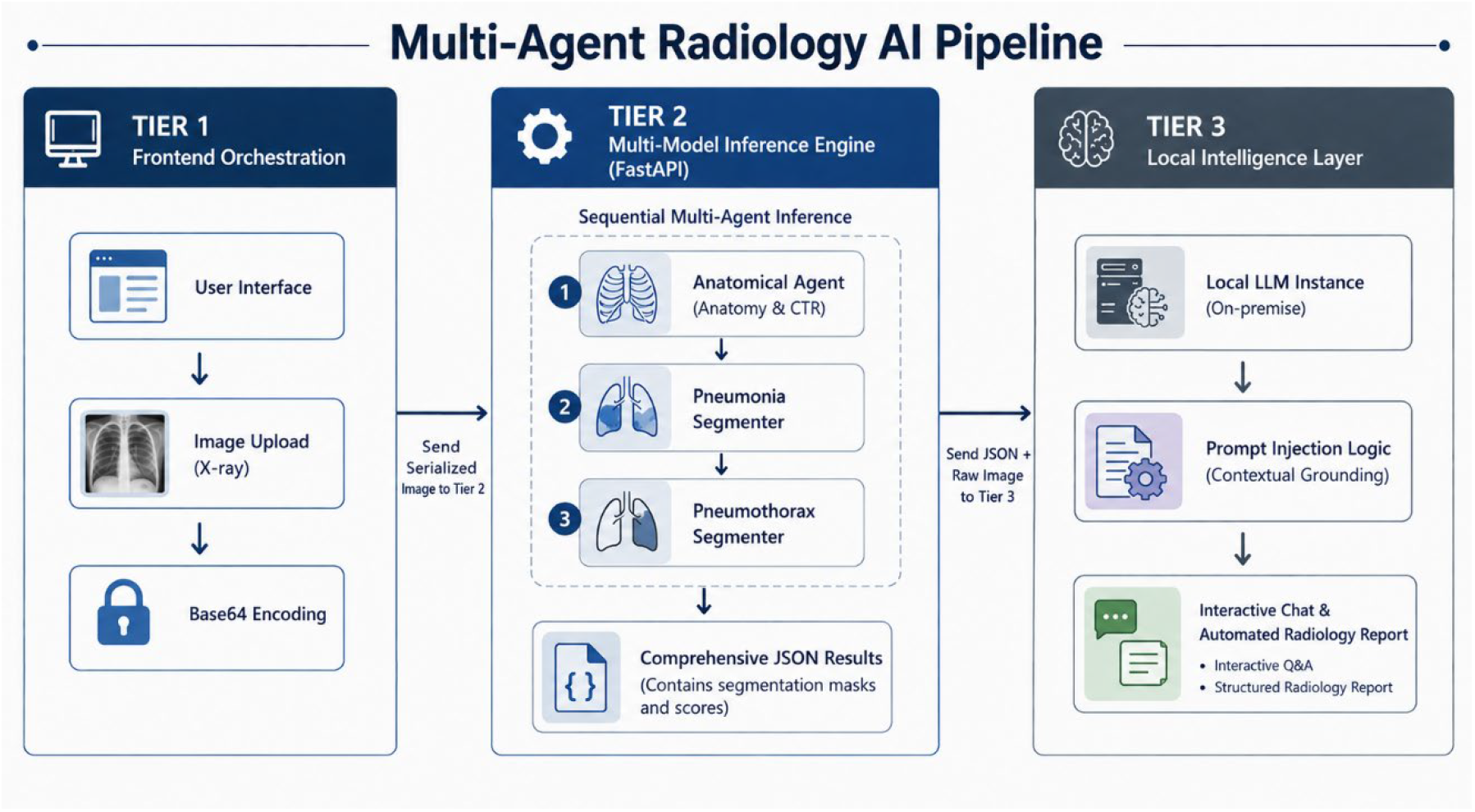
Architecture of the proposed multi-agent radiology AI pipeline integrating frontend orchestration, sequential multi-model inference, and a local intelligence layer for automated report generation and interactive clinical analysis.

**Figure 2:**
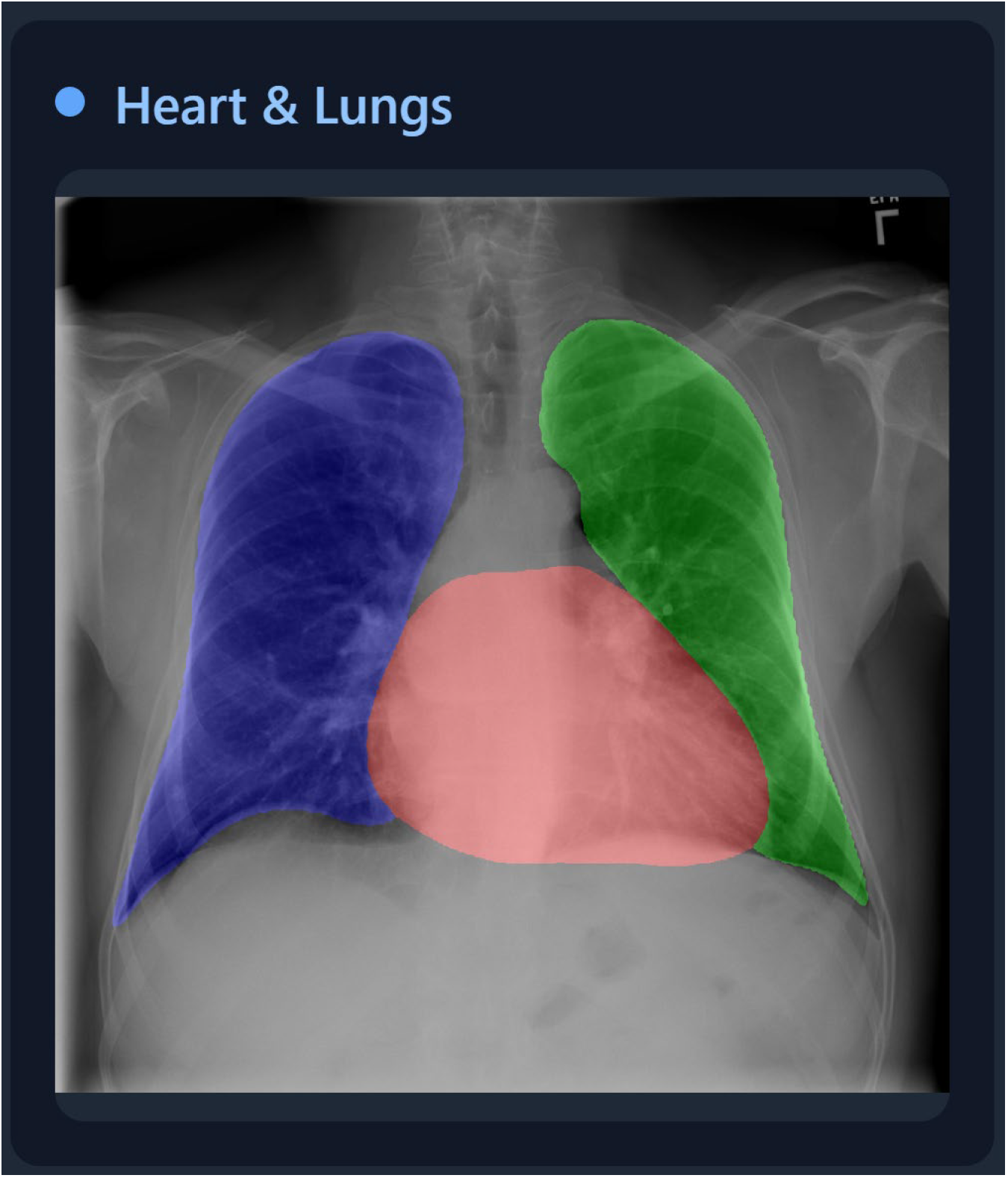
Anatomical segmentation of the lungs and cardiac silhouette on a sample from the NIH Chest X-ray dataset.

**Figure 3:**
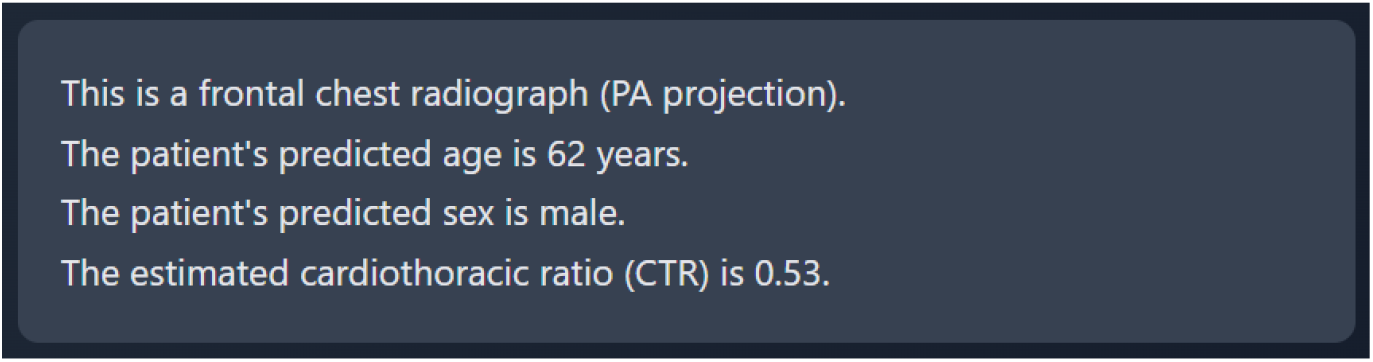
Automated demographic predictions and Cardiothoracic Ratio (CTR) calculated by the baseline agent.

**Figure 4:**
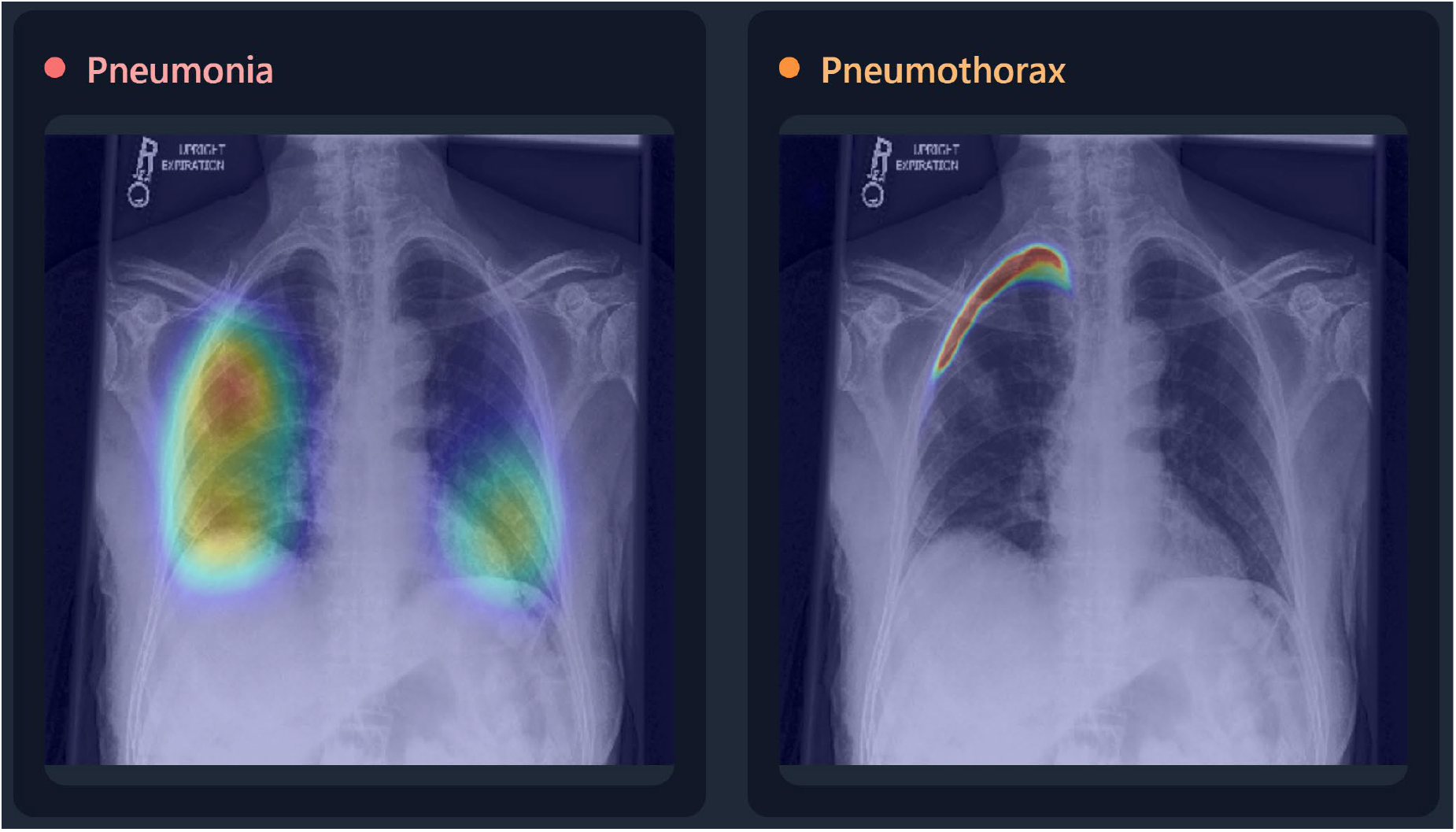
False-color segmentation masks generated by the pneumothorax and pneumonia agents, delineating the spatial extent of the detected pathology on a sample radiograph from a public Kaggle [34] dataset.

The first tier serves as the presentation and orchestration environment. This client-facing layer manages the initial ingestion of radiological data. When a user uploads a raw chest radiograph, the client application securely processes the file by converting it into a standardized, text-encoded format directly within the browser. This stateless approach eliminates the need for temporary cloud-based file storage, allowing the raw image data to be securely and directly transmitted as a serialized payload to the system’s internal routing gateways.

The second-tier functions as the core multi-model inference engine. Operating within a high-performance asynchronous framework, this backend layer is responsible for decoding the ingested image and orchestrating a robust computer vision pipeline. Rather than processing artificial intelligence tasks simultaneously, the engine enforces a strict, deterministic execution sequence. The visual data is first processed by an anatomical foundational model to extract demographic estimates and baseline anatomical metrics. Subsequently, the image is passed sequentially through specialized pathological segmentation networks designed to identify specific thoracic conditions. The inference engine then aggregates these disparate findings, including disease probability scores and generated pixel-level visual overlays, compiling them into a single, comprehensive structured data object.

The third and final tier is the intelligence and consultation layer, which houses the conversational vision-language model. To ensure absolute patient data privacy and adhere to strict medical data protection principles, this layer executes entirely on-premise without relying on external cloud application programming interfaces. The system utilizes LLaVA 7B [23] as the default multimodal large language model within this local environment. The central orchestration server synthesizes the multimodal inputs by constructing a highly structured textual prompt derived directly from the inference engine’s comprehensive data object. This structured clinical ground truth, paired with the original text-encoded radiograph, is then routed to the locally hosted language model. By simultaneously processing the raw visual data and the algorithmically derived diagnostic context, the system seamlessly transitions from deterministic image segmentation to dynamic, interactive clinical mentorship.

### 3.2. The Anatomical Baseline Agent

The multi-agent pipeline initiates its visual analysis through a foundational anatomical baseline agent, specifically utilizing the “ianpan/chest-x-ray-basic” [24] model. This component employs a multi-task deep learning architecture featuring a EfficientNetV2 [25] backbone model integrated with a U-Net [26] decoder for precise spatial segmentation, alongside a dedicated linear layer for metadata classification. The model was trained on a highly diverse corpus comprising 335,516 images from 96,385 patients, utilizing an 80 to 20 training and validation split derived from the CheXpert [27] and NIH Chest X-ray [28] datasets, with ground truth segmentation masks sourced from the CheXmask [29] dataset. This architecture processes the image to generate high-resolution, pixel-level segmentation masks through bilinear interpolation, isolating three primary anatomical regions with exceptional reliability. Performance metrics demonstrate a Dice similarity coefficient of 0.957 for the right lung, 0.948 for the left lung, and 0.943 for the cardiac silhouette.

Operating concurrently with the spatial segmentation, the agent utilizes its auxiliary linear classification head to execute end-to-end demographic and orientational predictions. To mitigate unwanted dataset bias, particularly because lateral radiographs are exclusive to the CheXpert [27] dataset, the view classifier was selectively trained solely on CheXpert [27] images while excluding NIH images from the loss function. This strategic training yields highly accurate orientational predictions, achieving a 99.42 percent accuracy rate in distinguishing between Anteroposterior, Posteroanterior, and Lateral views. This automatic view categorization is essential for contextualizing the spatial data and ensuring the validity of subsequent anatomical measurements. Furthermore, without relying on external clinical notes or hand-crafted heuristic rules, the model computes continuous scalar values to estimate patient demographics, achieving a mean absolute error of 5.25 years for age prediction and an Area Under the Curve of 0.999 for biological sex classification.

Leveraging the precise boundaries defined by the U-Net [26] segmentation masks, the agent automates the calculation of the Cardiothoracic Ratio. Rather than replicating the traditional manual measurement of distinct thoracic diameters, the system calculates a highly reproducible geometric proxy based on algorithmic bounding spans. It determines the maximum horizontal pixel extents of the segmented heart and divides this value by the combined maximum horizontal extents of the bilateral lung fields. To ensure clinical validity, this mathematical calculation is strictly gated by the view prediction module; it executes exclusively on frontal projections and automatically disables for lateral views to prevent erroneous anatomical assessments.

Finally, the agent synthesizes these extracted spatial geometries, demographic predictions, and calculated ratios into a unified, structured data payload. The visual segmentation overlays are encoded into text-safe formats alongside the numeric and categorical clinical metrics. This packaged anatomical baseline is subsequently injected directly into the conversational language model prompt. It is imperative to note that while this baseline agent provides a highly accurate structural foundation for the educational pipeline, the underlying model is deployed strictly for demonstration and research purposes. It has not been approved by any regulatory agency for clinical use, ensuring the system’s application remains firmly within the boundaries of a risk-free academic simulator rather than a definitive diagnostic tool.

### 3.3. Pathological Segmentation Agents

Following the successful extraction of the anatomical baseline, the multi-agent pipeline initiates a strict, deterministic sequence of pathological analyses. To maximize computational stability and isolate diagnostic reasoning, the system does not utilize parallel scheduling. Instead, the inference engine processes the decoded grayscale radiograph sequentially through dedicated disease-specific agents, completing the full pneumonia evaluation cycle before initiating the pneumothorax assessment.

The primary pathological agent dedicated to pneumonia detection (utilizing the *ianpan/pneumonia-cxr* [30] model) employs a multi-task deep learning architecture consisting of an EfficientNetV2-S backbone integrated with a U-Net decoder and a linear classification head. This architecture was robustly trained on the RSNA Pneumonia Detection Challenge [31] and the SIIM-FISABIO-RSNA COVID-19 Detection [32] datasets. Notably, to facilitate dense pixel-level training, the original bounding box annotations from these datasets were algorithmically transformed into continuous ellipsoid segmentation masks. On independent holdout test sets, this model demonstrated robust diagnostic capabilities, achieving a combined Area Under the Curve of 0.900. Following the pneumonia evaluation, the pipeline invokes the secondary pathological agent dedicated exclusively to pneumothorax detection (utilizing the *ianpan/pneumothorax-cxr* [33] model). Operating on an identical structural framework, this model leverages an EfficientNetV2-S [25] encoder paired with a 5-block U-Net-style [26] decoder to process the single-channel, 512x512 resolution input. Without relying on generic classification networks, this custom architecture produces both a dense spatial map and an independent scalar disease score specifically calibrated for pneumothorax detection.

During the visual extraction phase, neither pathological agent relies on traditional bounding box coordinates. Instead, each model independently preprocesses the image and generates a dense single-channel mask tensor. These spatial activations are resized to the original high-resolution image dimensions using bilinear interpolation, scaled appropriately, and translated through a false color colormap. This mathematical process produces precise, pixel-level visual overlays that emphasize the exact location and extent of the pathology, which are subsequently blended onto the RGB rendering of the original radiograph. With spatial segmentation, the agents perform mathematical binary classification by extracting a continuous scalar probability value from their respective linear heads. The backend enforces a rigid thresholding mechanism to categorize the disease state. A pathology is classified as detected exclusively if the scalar probability is strictly greater than 0.5; a score exactly equal to this threshold defaults to a negative classification. Furthermore, the system computes a calibrated confidence metric for each prediction, calculated as the absolute mathematical distance from decision ambiguity. This is defined as the maximum value between the predicted probability and its inverse, ensuring the reported confidence score accurately reflects the certainty of the binary decision rather than functioning as a raw probability output.

Upon completion of these sequential forward passes, the extracted pathological findings are dynamically integrated into the unified diagnostic payload. This structured data object systematically catalogs the continuous probabilities, the discrete Boolean detection flags, and the calculated confidence scores for both targeted diseases. Simultaneously, the false-color segmentation overlays are securely text-encoded and appended alongside the baseline anatomical maps. By packaging these highly specific pathological outputs with the underlying anatomical geometry, the inference engine constructs a comprehensive, mathematically grounded context that tightly governs the subsequent deductive reasoning of the large language model.

### 3.4. Multi-Model Data Fusion and Contextual Linearization

A fundamental challenge in multi-agent artificial intelligence is effectively transferring the mathematical outputs of deterministic computer vision models into the generative reasoning space of a language model. Rather than transmitting raw data structures directly to the consultation layer, the system employs a selective linearization process. The comprehensive diagnostic payload, which contains baseline anatomical metrics and pathological probabilities, is algorithmically transformed into a highly structured plain-text summary.

During interactive consultation, this fusion process selectively extracts the narrative demographic information, the binary disease classifications, and their associated confidence percentages. Furthermore, the presence of spatial segmentation maps is qualitatively noted within the text, ensuring the language model is aware of the visual evidence without requiring the computationally expensive embedding of secondary image files. For automated report generation, this fusion process is significantly expanded. The server synthesizes the quantitative probabilities and anatomical metrics into formal clinical subsections, preparing a structured prose foundation before language generation begins. Ultimately, this linearized textual context is paired exclusively with the raw, text-encoded original radiograph, creating a multiplexed visual and linguistic payload tailored specifically for the local multimodal model.

### 3.5. LLM Integration and Epistemic Guardrails

To mitigate the severe hallucination risks inherent in multimodal models, the system implements a strict, prompt-based grounding architecture utilizing LLaVA 7B [23] as the primary inference engine. This model executes entirely within a localized environment via the Ollama [35] framework, where the linearized clinical summary is encapsulated within rigid instructional guardrails. These parameters dictate strict formatting constraints, limiting responses to concise paragraphs while explicitly forbidding structural deviations like bulleted lists. More importantly, the system enforces a strict epistemic boundary. The language model is explicitly instructed to base its clinical interpretations solely on the provided algorithmic analysis, to unequivocally refuse the fabrication of absent findings, and to mandate professional medical consultation.

A critical safety feature of this integration is the prerequisite protocol, designed to prevent unanchored visual interpretation. If a user attempts to initiate a clinical dialogue before executing the foundational visual analysis, the orchestration layer intercepts the request. Instead of allowing the multimodal model to blindly interpret the radiograph, the system dynamically substitutes the system instructions, steering the language model to politely refuse the query and direct the user to complete the multi-model analysis first. In the context of automated report generation, this prerequisite is enforced as a hard architectural block, terminating the request entirely if the mathematical ground truth is absent.

Additionally, the system employs post-generation textual filtering mechanisms to eliminate any residual model-specific artifacts or vendor nomenclature from the generated prose. By combining instructional prompt contracts, dynamic request interception, and post-generation sanitation, the system effectively constrains the generative capabilities of the local model, ensuring it functions as an objective educational simulator strictly tethered to the deterministic visual evidence.

### 3.6. Automated Structured Report Generation

Beyond interactive consultation, the system leverages the multi-agent pipeline to function as an automated medical scribe, transforming quantitative visual findings into formal, structured radiology reports. This capability is governed by a secondary, highly specialized instructional template that imposes strict stylistic and structural requirements on the generative model. Rather than utilizing downstream parsers or external schema validators, the system enforces document structure entirely through prompt engineering. The template explicitly mandates the sequential inclusion of standard radiological headers, specifically requiring sections for Patient Information, Clinical Technique, Narrative Findings, and a final Numbered Impression.

To ensure the generated text accurately reflects the algorithmic analysis, the orchestration layer performs a dynamic synthesis of the available quantitative data before generation begins. The categorical disease classifications, continuous probability scores, and calculated baseline metrics such as the Cardiothoracic Ratio are mathematically normalized and systematically interpolated into the instructional prompt. Instead of passing the raw data objects to the model, the system linearizes these values into thematic integration paragraphs. This method forces the language model to synthesize the statistical results with its visual perception, ensuring the generated narrative findings are heavily anchored to the deterministic outputs of the segmentation agents.

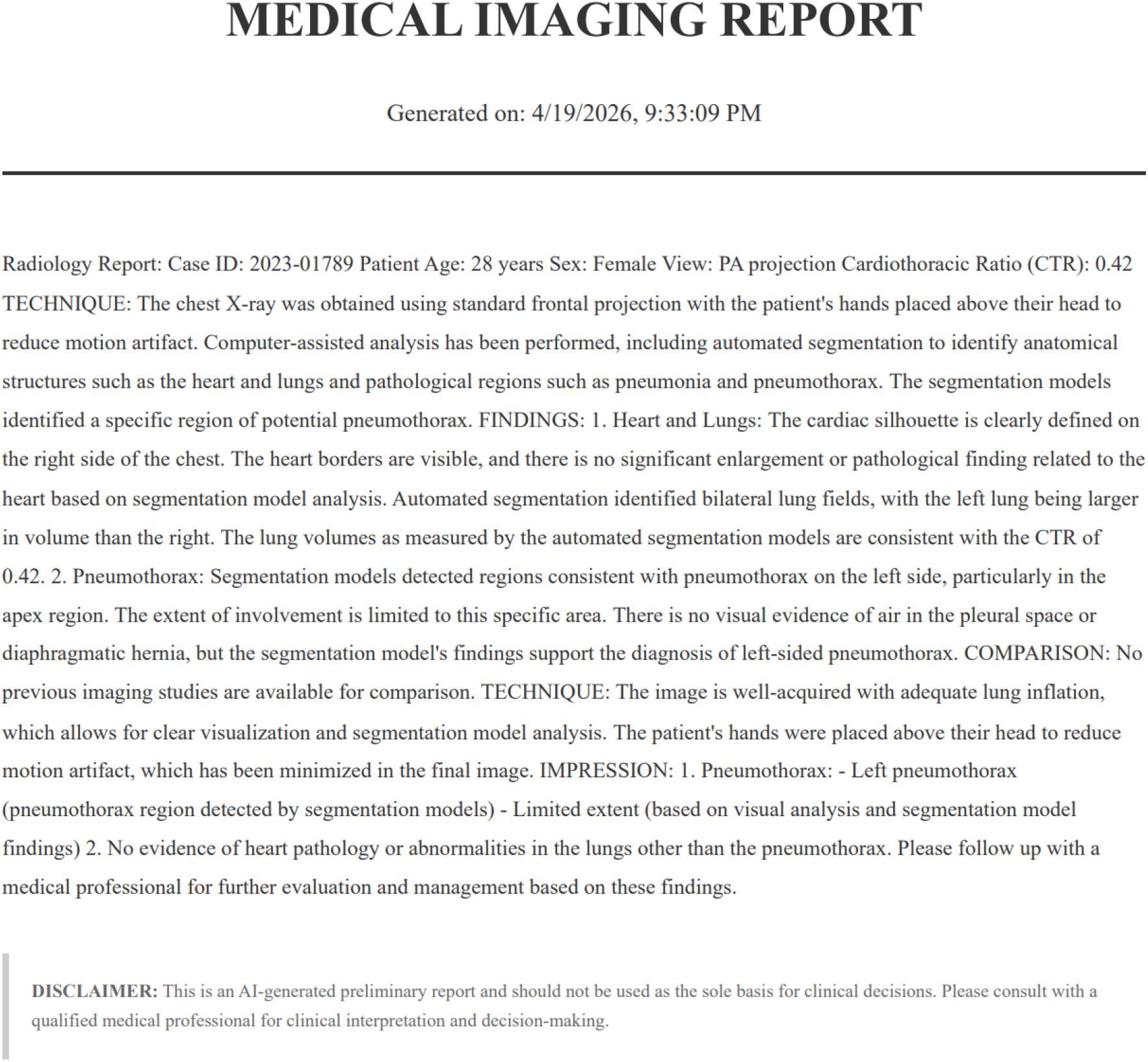

Crucially, the system imposes severe constraints on the clinical tone and nomenclature used within the report. The prompt provides explicit stock phrasing to ensure the model adopts professional radiological dictation standards, requiring the attribution of findings to neutral terms such as computer-assisted analysis or automated segmentation models. Furthermore, to maintain the professional integrity of the simulated environment, the model is explicitly forbidden from referencing its own architectural identity or underlying vendor technologies within the text. To reinforce these epistemic constraints, the pipeline executes a post-generation sanitization protocol before presenting the final report to the user. The orchestration server performs a systematic string evaluation of the generated output, aggressively identifying and stripping any residual vendor names, model identifiers, or colloquial artificial intelligence terminology. Through this combination of strict prompt templates, dynamic data interpolation, and post-generation filtering, the system successfully translates raw multi-model visual data into a cohesive, formally structured clinical document, maximizing the educational utility of the risk-free training environment.

## 4. System Implementation and Interactive Demonstration

To evaluate the practical efficacy of sequential multi-agent architecture, the system was deployed in a simulated educational environment. The following clinical interaction scenarios demonstrate how the computational pipeline translates into a safe, interactive mentorship experience for medical trainees.

### 4.1. Scenario 1: Prerequisite Protocol Enforcement

The primary vulnerability of multimodal large language models is their tendency to fabricate clinical findings when prompted with unanalyzed visual data. To demonstrate the system’s epistemic guardrails, a test scenario was initiated where the user uploads a chest radiograph and immediately asks the system to identify visible pathologies. As depicted in Figure 5, rather than attempting a blind visual interpretation, the orchestration layer successfully intercepts the query. The system explicitly refuses the prompt, informing the user that it cannot interpret raw medical images independently. It then actively directs the trainee to execute the comprehensive multi-model analysis before proceeding. This interaction validates the prerequisite protocol, proving that the system successfully prioritizes mathematical grounding over conversational compliance.

**Figure 5:**
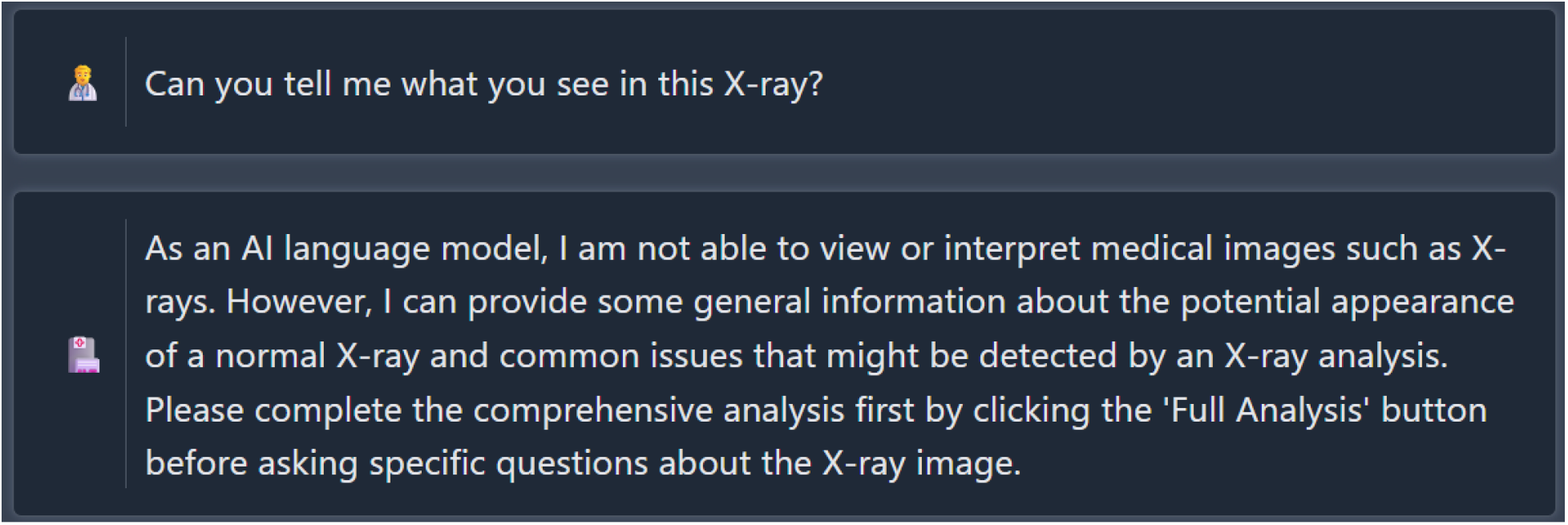
Demonstration of the prerequisite protocol. The system actively intercepts a clinical query and refuses to hallucinate visual findings without prior multi-model quantitative analysis, directing the user to complete the data extraction phase.

### 4.2. Scenario 2: Anatomical Education and Grounding

Once the visual baseline is established, the system functions as an anatomical tutor. In this scenario, the user queries the significance of the automatically calculated Cardiothoracic Ratio of 0.51. As illustrated in Figure 6, because the language model has been injected with the structured quantitative payload, it does not generate a generic definition. Instead, artificial intelligence contextualizes the specific mathematical output, explaining that a ratio of 0.51 indicates the heart occupies approximately half of the thoracic cavity, which falls within normal physiological limits for the predicted demographic. This demonstrates the system’s capacity to bridge the gap between abstract algorithmic outputs and practical anatomical education, allowing trainees to understand the clinical meaning behind the extracted baseline metrics.

**Figure 6:**
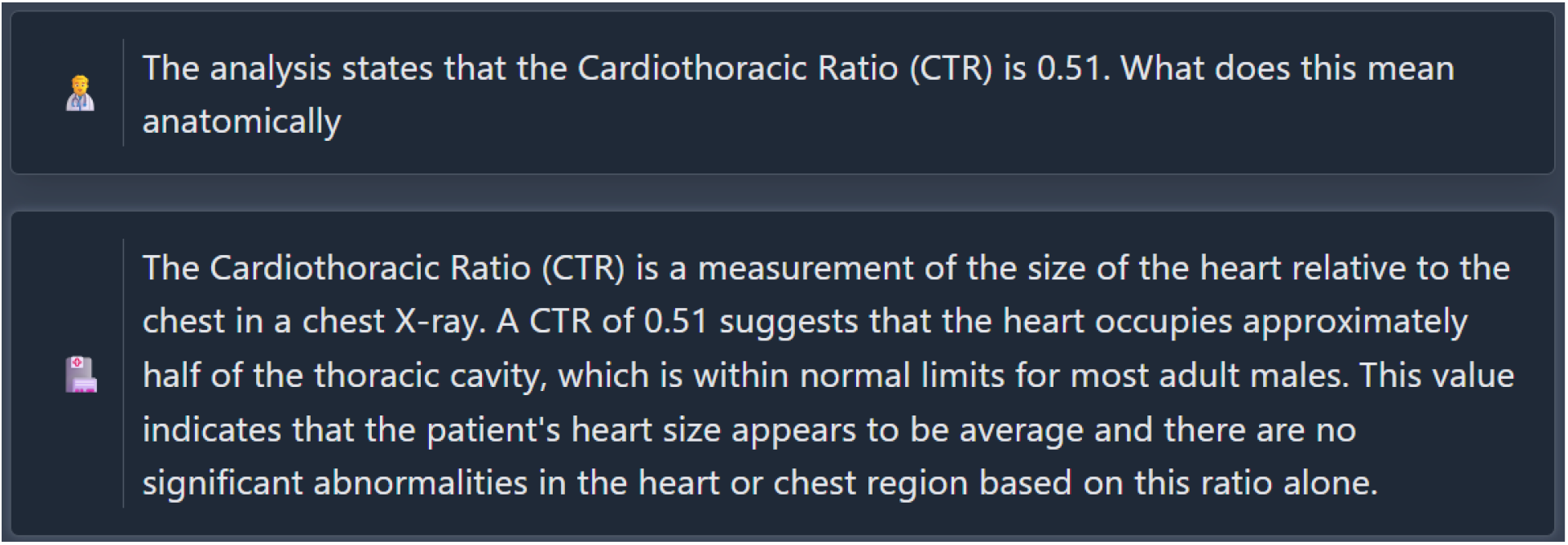
The system contextualizes the Cardiothoracic Ratio (0.51), providing an anatomically grounded educational explanation tailored to the patient demographic.

### 4.3. Scenario 3: Pathology Explanation and Safety Guardrails

The final scenario evaluates the system’s ability to explain pathological findings without violating medical safety constraints. As depicted in Figure 7, the user asks the system to describe the visual characteristics of an opacity identified by the pneumonia segmentation agent. Relying strictly on the fused quantitative payload, the system confirms the location of the opacity in the right lung and references the precise 84 percent confidence score generated by the underlying deep learning model. It explains that the density corresponds to potential pulmonary consolidation. Crucially, the system concludes the interaction by automatically appending a safety disclaimer, reminding the trainee that visual analysis requires further medical assessment for an accurate diagnosis. This interaction confirms that the system can successfully integrate advanced algorithmic metrics into a conversational framework while maintaining strict medico-legal boundaries.

**Figure 7:**
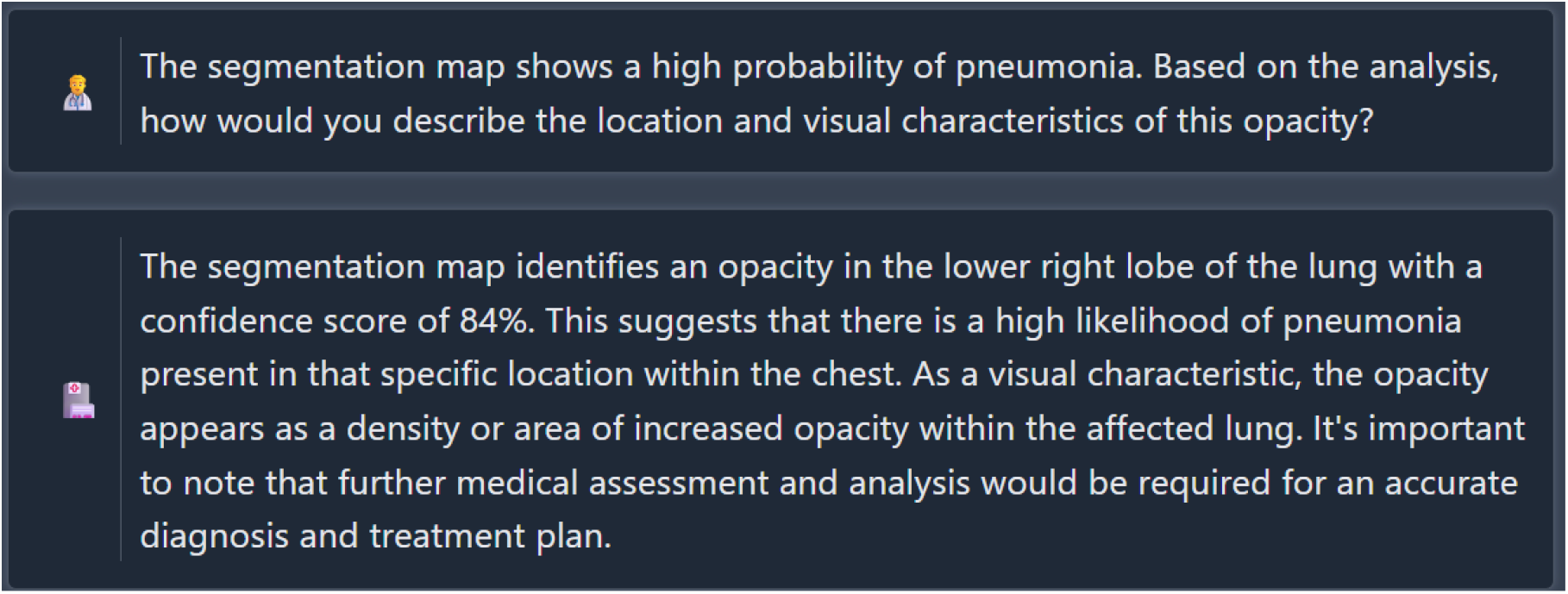
Pathology explanation demonstrating safety guardrails. The system contextualizes the 84% confidence score while enforcing strict medical disclaimers.

## 5. Limitations and Future Directions

While the sequential multi-agent architecture successfully mitigates multimodal hallucinations, the framework possesses inherent limitations. Primarily, the system’s diagnostic accuracy is strictly bounded by the generalizability of its underlying computer vision agents. Because the foundational deep learning models were trained on specific adult datasets, such as the RSNA and NIH collections, their performance may demonstrate variance when applied to pediatric cases or divergent demographic populations. Furthermore, the deliberate architectural decoupling of visual extraction and language generation introduces a structural text-bottleneck. By forcing the language model to rely exclusively on the intermediate JSON payload to ensure epistemic safety, the system becomes completely dependent on the upstream segmentation accuracy. If a specialized agent fails to detect a specific lesion during the visual extraction phase, the language model is mathematically blind to its presence, regardless of its visibility in the raw radiograph.

Subsequent development phases will focus on validating the educational utility and expanding the diagnostic breadth of the architecture. A primary objective is the implementation of structured clinical pilot testing to quantitatively assess the pedagogical impact of the artificial intelligence tutor on the diagnostic reasoning of medical students. Architecturally, the highly modular nature of the inference engine facilitates the seamless integration of expanded pathology detection. Future iterations will incorporate additional plug-and-play segmentation agents capable of identifying a broader spectrum of thoracic conditions, such as pleural effusion and cardiomegaly. Finally, to elevate the system’s reasoning capabilities, future research will focus on integrating advanced data retrieval mechanisms. By bringing the existing architecture with Medical Knowledge Graphs and Retrieval-Augmented Generation frameworks together, the language model will be able to cross-reference its extracted visual findings with dynamically retrieved, evidence-based medical literature. This will evolve the system from an isolated anatomical tutor into a comprehensive, continuously updating clinical simulator.

## 6. Conclusion

This research presents a novel multi-agent architecture designed to transform traditional black-box radiology artificial intelligence into an interactive and explainable educational tool. By decoupling deterministic image analysis from generative linguistic consultation, the system successfully addresses the critical challenge of hallucinations in medical large language models. The integration of specialized segmentation agents provides a verified mathematical ground truth, ensuring that the conversational tutor remains strictly anchored to the visual evidence.

Through the implementation of rigorous epistemic guardrails and a sequential inference pipeline, the framework offers a safe, infinitely available environment for medical trainees to develop their diagnostic reasoning skills. The ability to generate automated, structured reports while maintaining a dynamic consultative dialogue proves that artificial intelligence can function not merely as a diagnostic endpoint, but as a sophisticated clinical mentor. Ultimately, this work establishes a new paradigm for consultative artificial intelligence in healthcare, demonstrating that the strategic synthesis of high-precision computer vision and local vision-language models can bridge the gap between raw data extraction and meaningful clinical interpretation. As artificial intelligence continues to integrate into medical informatics, such grounded and explainable systems will be essential in shaping the next generation of digitally empowered medical professionals.

## Data Availability

All data produced are available online at
https://www.kaggle.com/datasets/nih-chest-xrays/data/data
https://stanfordmlgroup.github.io/competitions/chexpert/

https://stanfordmlgroup.github.io/competitions/chexpert/

## Notes

### Competing Interest Statement

The authors have declared no competing interest.

## References

[1] R. Lorenzo-Alvarez, J. Pavia-Molina, and F. Sendra-Portero, “Exploring the Potential of Undergraduate Radiology Education in the Virtual World Second Life with First-cycle and Second-cycle Medical Students,” Acad. Radiol., vol. 25, no. 8, pp. 1087–1096, Aug. 2018, doi: 10.1016/j.acra.2018.02.026.

[2] S. Prathyunman and S. P. Kasthuri Arachchi, “Interpretation of Chest X-rays using Deep Learning Techniques for Early Diagnosis of Tuberculosis,” in 2025 5th International Conference on Advanced Research in Computing (ICARC), Belihuloya, Sri Lanka: IEEE, Feb. 2025, pp. 1–6. doi: 10.1109/ICARC64760.2025.10962936.

[3] W. Pan, S. Li, B. Tian, Y. Zheng, and H. Wang, “Artificial intelligence in medical education for pulmonary nodule management: a narrative review,” Transl. Lung Cancer Res., vol. 14, no. 9, pp. 4068–4077, Sep. 2025, doi: 10.21037/tlcr-2025-313.

[4] K. Shaw, M. Henning, and C. Webster, “Artificial Intelligence in Medical Education: A Scoping Review of the Evidence for Efficacy and Future Directions,” 2024, SSRN. doi: 10.2139/ssrn.4673012.

[5] A. B. Das, S. K. Sakib, and S. Ahmed, “Trustworthy Medical Imaging with Large Language Models: A Study of Hallucinations Across Modalities”.

[6] S. Salehi, Y. Singh, K. K. Horst, Q. A. Hathaway, and B. J. Erickson, “Agentic AI and Large Language Models in Radiology: Opportunities and Hallucination Challenges,” Bioengineering, vol. 12, no. 12, p. 1303, Nov. 2025, doi: 10.3390/bioengineering12121303.

[7] M. Haupt and M. H. Maurer, “Explainable artificial intelligence in radiology: Methods, clinical applications, limitations, and future directions,” Eur. J. Radiol. Artif. Intell., vol. 6, p. 100098, Jun. 2026, doi: 10.1016/j.ejrai.2026.100098.

[8] M. U. Alam, J. Hollmén, J. R. Baldvinsson, and R. Rahmani, “SHAMSUL: Systematic Holistic Analysis to investigate Medical Significance Utilizing Local interpretability methods in deep learning for chest radiography pathology prediction,” Nord. Mach. Intell., vol. 3, no. 1, pp. 27–47, Aug. 2024, doi: 10.5617/nmi.10471.

[9] L. Pawar and D. S. Patil, “Explainable AI Techniques for Lung Disease Detection: Comparative Foundations, Experimental Evaluation, Challenges, and Future Directions”.

[10] Z. Qiu, “Enhancing Visual Interpretability in Computer-Assisted Radiological Diagnosis: Deep Learning Approaches for Chest X-Ray Analysis”.

[11] E. Koutoulakis, E. Trivizakis, E. Markodimitrakis, S. Agelaki, M. Tsiknakis, and K. Marias, “A critical review of explainable deep learning in lung cancer diagnosis,” Artif. Intell. Rev., vol. 59, no. 1, p. 28, Dec. 2025, doi: 10.1007/s10462-025-11445-x.

[12] D. Chen, A. Ayoob, T. S. Desser, and A. Khurana, “Review of Learning Tools for Effective Radiology Education During the COVID-19 Era,” Acad. Radiol., vol. 29, no. 1, pp. 129–136, Jan. 2022, doi: 10.1016/j.acra.2021.10.006.

[13] S. W. T. Wade, G. M. Velan, N. Tedla, N. Briggs, and M. Moscova, “What works in radiology education for medical students: a systematic review and meta-analysis,” BMC Med. Educ., vol. 24, no. 1, p. 51, Jan. 2024, doi: 10.1186/s12909-023-04981-z.

[14] S. Wade, “Undergraduate Medical Student Education in Radiology: The Role of eLearning,” UNSW Sydney, 2024. doi: 10.26190/UNSWORKS/25488.

[15] I. Altun, O. Turan, and O. Awan, “Revolutionizing radiology education: exploring innovative teaching methods,” Abdom. Radiol., vol. 50, no. 12, pp. 6225–6234, Jun. 2025, doi: 10.1007/s00261-025-05010-x.

[16] D. A. Yip, D. G. Craig, D. J. Cortes-Ramirez, N. White, K. Shaw, and S. Reddy, “Patient-centric radiology: Utilising large language models (LLMs) to improve patient communication and education”.

[17] M. S. Von Der Stück et al., “Visual Large Language Models in Radiology: A Systematic Multimodel Evaluation of Diagnostic Accuracy and Hallucinations,” Life, vol. 16, no. 1, p. 66, Jan. 2026, doi: 10.3390/life16010066.

[18] A. B. Das, S. K. Sakib, and S. Ahmed, “Trustworthy Medical Imaging with Large Language Models: A Study of Hallucinations Across Modalities”.

[19] Y. Kim et al., “Medical Hallucinations in Foundation Models and Their Impact on Healthcare,” 2025, arXiv. doi: 10.48550/ARXIV.2503.05777.

[20] E. Asgari et al., “A Framework to Assess Clinical Safety and Hallucination Rates of LLMs for Medical Text Summarisation,” Sep. 13, 2024, Health Informatics. doi: 10.1101/2024.09.12.24313556.

[21] E. Nasarian, A. Neog, K.-L. Tsui, and N. HosseiniChimeh, “CareGuardAI: Context-Aware Multi-Agent Guardrails for Clinical Safety & Hallucination Mitigation in Patient-Facing LLMs”.

[22] D. Wang et al., “Enhancing Large Language Models for Improved Accuracy and Safety in Medical Question Answering: Comparative Study,” JMIR Med. Educ., vol. 11, p. e70190, Dec. 2025, doi: 10.2196/70190.

[23] H. Liu, C. Li, Q. Wu, and Y. J. Lee, “Visual Instruction Tuning,” in Advances in Neural Information Processing Systems, A. Oh, T. Naumann, A. Globerson, K. Saenko, M. Hardt, and S. Levine, Eds., Curran Associates, Inc., 2023, pp. 34892–34916. [Online]. Available: https://proceedings.neurips.cc/paper_files/paper/2023/file/6dcf277ea32ce3288914faf369fe6de0-Paper-Conference.pdf

[24] “ianpan/chest-x-ray-basic · Hugging Face.” Accessed: May 10, 2026. [Online]. Available: https://huggingface.co/ianpan/chest-x-ray-basic

[25] M. Tan and Q. V. Le, “EfficientNetV2: Smaller Models and Faster Training”.

[26] O. Ronneberger, P. Fischer, and T. Brox, “U-Net: Convolutional Networks for Biomedical Image Segmentation,” in Medical Image Computing and Computer-Assisted Intervention – MICCAI 2015, vol. 9351, N. Navab, J. Hornegger, W. M. Wells, and A. F. Frangi, Eds., in Lecture Notes in Computer Science, vol. 9351., Cham: Springer International Publishing, 2015, pp. 234–241. doi: 10.1007/978-3-319-24574-4_28.

[27] J. Irvin et al., “CheXpert: A Large Chest Radiograph Dataset with Uncertainty Labels and Expert Comparison,” Proc. AAAI Conf. Artif. Intell., vol. 33, no. 01, pp. 590–597, Jul. 2019, doi: 10.1609/aaai.v33i01.3301590.

[28] X. Wang, Y. Peng, L. Lu, Z. Lu, M. Bagheri, and R. M. Summers, “ChestX-ray8: Hospital-Scale Chest X-Ray Database and Benchmarks on Weakly-Supervised Classification and Localization of Common Thorax Diseases”.

[29] N. Gaggion et al., “CheXmask: a large-scale dataset of anatomical segmentation masks for multi-center chest x-ray images,” Sci. Data, vol. 11, no. 1, p. 511, May 2024, doi: 10.1038/s41597-024-03358-1.

[30] “ianpan/pneumonia-cxr · Hugging Face.” Accessed: May 10, 2026. [Online]. Available: https://huggingface.co/ianpan/pneumonia-cxr

[31] M. Anouk Stein et al., “RSNA Pneumonia Detection Challenge.” 2018. [Online]. Available: https://kaggle.com/competitions/rsna-pneumonia-detection-challenge

[32] A. Kemp et al., “SIIM-FISABIO-RSNA COVID-19 Detection.” 2021. [Online]. Available: https://kaggle.com/competitions/siim-covid19-detection

[33] “ianpan/pneumothorax-cxr · Hugging Face.” Accessed: May 10, 2026. [Online]. Available: https://huggingface.co/ianpan/pneumothorax-cxr

[34] “Pneumothorax.” Accessed: May 11, 2026. [Online]. Available: https://www.kaggle.com/datasets/adnanenasser/pneumothorax

[35] “Ollama.” Accessed: May 11, 2026. [Online]. Available: https://ollama.com

